# Potential five-year life years associated with stage IV reductions in NHS Galleri

**DOI:** 10.64898/2026.09.21.26363618

**Authors:** Natallia Starasvetskay, Gilberto de Lima Lopes

**Author notes:** Corresponding author: Gilberto de Lima Lopes, Jr., MD, MBA. 1120 NW 14^th^ street, suite 650L, Miami, FL 33136.

## Abstract

**Background:** NHS-Galleri did not reach its prespecified primary endpoint of a reduction in combined stage III and IV cases, but reported fewer stage IV cancers after annual multi-cancer early detection screening. We estimated the potential five-year life-years associated with the cancer-specific stage IV differences observed during the two incident screening rounds.

**Methods:** We used intervention and control stage IV counts for ten prespecified cancers reported in the NHS-Galleri ASCO 2026 presentation. For each cancer, the randomized-arm count difference was multiplied by the difference in five-year restricted mean survival time between stage III and IV, estimated from England stage-specific five-year net survival using a constant-hazard fit. Adverse cancer-specific differences remained in the model. Results were annualized across the two incident rounds and scaled per million people screened. Sampling intervals used an independent Poisson approximation for arm-specific counts.

**Findings:** Across the ten evaluable cancers, the intervention and control arms had 180 and 239 stage IV diagnoses during the incident rounds. The conditional projection was 66.3 potential life-years over five years in the trial intervention cohort, equivalent to 466 per million people screened annually (95% sampling interval 190-743). For a population of 10 million, the corresponding projection was 4,664 annually (1,896-7,432). Colorectal, lung, and esophageal cancers accounted for 79% of the net projection. Pancreatic, ovarian, and lymphoid cancers contributed negatively because their observed stage IV counts favored control.

**Interpretation:** If the observed incident-round reduction in stage IV diagnoses represents true downstaging to stage III, it could produce meaningful population health gains. This conditional estimate is not evidence of mortality reduction. Longer follow-up remains necessary to distinguish benefit from chance, lead-time and length biases, overdiagnosis, and differences in tumor biology.

## Introduction

The NHS-Galleri trial randomized 142,924 asymptomatic adults to annual multi-cancer early detection testing plus standard care or standard care alone. After three screening rounds, the trial did not meet its primary endpoint for combined stage III and IV cancers among 12 prespecified cancer types (706 vs 688; incidence rate ratio 1.03, 95% CI 0.92-1.14). It reported fewer stage IV cancers overall (342 vs 397; incidence rate ratio 0.86, 95% CI 0.744-0.998), with progressively larger differences in the second and third screening rounds.

The clinical meaning of a stage IV reduction is uncertain because stage shift is a surrogate rather than a mortality endpoint. Nevertheless, stage IV and stage III cancers have substantially different survival across several tumor types. We developed a conditional projection using only randomized incident-round stage IV differences. We did not add the observed stage I-II excess, because doing so could count the same stage redistribution twice and because the reported cancer-specific early-stage counts cover a different three-round observation window.

## Methods

### Trial inputs

Cancer-specific stage IV counts were transcribed from slide 13 of the NHS-Galleri ASCO 2026 presentation. These counts cover the second and third annual screening rounds, both described as incident rounds. Ten cancers had reported counts and corresponding stage III and IV five-year net survival estimates: bladder, liver or bile duct, esophagus, head and neck, colorectum, lung, stomach, lymphoma, ovary, and pancreas. Anal cancer and myeloma were not modeled because the presentation did not report their stage-specific survival values. Their aggregate omitted stage IV difference was one event for anal cancer and zero for myeloma.

### Conditional projection

For cancer “*c”*, the net stage IV difference was defined as the control count minus the intervention count. Positive values therefore represented fewer stage IV diagnoses with screening, whereas negative values represented more. No cancer was excluded or reset to zero on the basis of effect direction.

Five-year restricted mean survival time was estimated from five-year net survival S using a constant-hazard exponential model: *RMST(5) = (1-S)/[-ln(S)/5]*. The potential life-years for each cancer equaled the net stage IV difference multiplied by RMST(5) for stage III minus RMST(5) for stage IV. This assigns each net stage IV diagnosis avoided to stage III and should be interpreted as a conditional stage-downshifting scenario.

The total was divided by the intervention-arm denominator of 71,122 and by two incident screening rounds, then multiplied by one million. Approximate sampling variance was the sum across cancers of (intervention count + control count) multiplied by the squared RMST difference. We report normal-approximation 95% sampling intervals. These intervals incorporate count uncertainty but not uncertainty in stage-specific survival or structural assumptions.

## Results

The ten evaluable cancers had 180 stage IV diagnoses in the intervention arm and 239 in control during the two incident rounds. The weighted difference corresponded to 66.3 conditional potential life-years over the five-year horizon in the trial intervention cohort. Annualized, this was 466 potential life-years per million people screened (95% sampling interval 190-743), or 4,664 per 10 million (1,896-7,432).

**Table 1.** Cancer-specific randomized counts and conditional annual contribution. Negative values remain in the total.

| Cancer | Intervention stage IV | Control stage IV | Annual potential LY per million |
| --- | --- | --- | --- |
| Bladder | 1 | 5 | 37.2 |
| Liver or bile duct | 4 | 14 | 58.0 |
| Oesophagus | 9 | 21 | 84.8 |
| Head and neck | 10 | 16 | 22.4 |
| Colorectum | 21 | 32 | 156.5 |
| Lung | 50 | 73 | 127.3 |
| Stomach | 6 | 8 | 16.2 |
| Lymphoma | 38 | 37 | -1.4 |
| Ovary | 8 | 7 | -4.9 |
| Pancreas | 33 | 26 | -29.6 |

Colorectal cancer contributed 156.5 life-years per million annually, lung 127.3, and oesophageal cancer 84.8. Together they accounted for 79% of the net projection. Pancreatic cancer contributed - 29.6 because the incident-round stage IV count was higher in the intervention arm (33 vs 26). Ovarian cancer and lymphoma also made small negative contributions. These are descriptive randomized differences, not cancer-specific efficacy estimates.

## Discussion

This simplified model produces a substantially smaller and more defensible projection than an approach that adds stage IV reductions to the stage I-II excess. The latter would combine cancer-specific observations measured over different windows and might count the same stage redistribution twice. Restricting the model to incident-round stage IV differences avoids that overlap and retains adverse cancer-specific observations.

The projection remains conditional. Stage-specific survival differences are not causal treatment effects. Lead-time bias, length bias, overdiagnosis, competing mortality, tumor aggressiveness, and treatment selection can all make earlier-stage survival appear better even when screening does not extend life. The model therefore quantifies the potential magnitude of benefit if the randomized stage IV difference proves to represent true downstaging to stage III. A sensitivity interpretation is linear: if only half of the modeled survival difference is realized, the projection is 233 life-years per million annually; at one quarter, it is 117.

PATHFINDER 2 provides complementary evidence about test performance and diagnostic pathways but cannot estimate stage shift or mortality benefit because it is single-arm. It reported 69.8% episode sensitivity for a prespecified group of 12 cancers responsible for approximately two-thirds of US cancer deaths, and 53% of newly detected primary cancers were stage I-II. Those findings support feasibility and detection performance, not the causal assumptions in this projection.

Cancer-specific findings should be interpreted cautiously. The event counts are small and were not powered for separate efficacy conclusions. The adverse pancreatic and ovarian point estimates do not establish harm or absence of benefit, just as the favorable bladder point estimate does not establish an 80% cancer-specific effect. The randomized aggregate result and subsequent mortality follow-up should remain primary.

The principal policy implication is narrow. A stage IV endpoint may contain clinically relevant information that a combined stage III-IV endpoint obscures, but it cannot substitute indefinitely for mortality. Future MCED trials should prespecify stage IV incidence, cancer-specific effects where adequately powered, interval and emergency presentations, diagnostic burden, quality of life, and cancer-specific and all-cause mortality.

## Limitations

The analysis relies on detailed data, but which was presentation-level data, still pending peer-reviewed publication. It assumes that each net stage IV diagnosis avoided becomes stage III, uses a constant-hazard fit based only on five-year net survival, and excludes uncertainty in survival estimates. It does not model competing mortality, costs, quality adjustment, diagnostic harms, adherence, or repeated lifetime screening. Anal cancer was omitted because the required survival contrast was unavailable on the slide, leaving one favorable stage IV event unmodeled. National extrapolation assumes the trial-arm rate applies at scale.

## Conclusion

The incident-round stage IV differences in NHS-Galleri correspond to approximately 466 conditional potential life-years per million people screened annually over a five-year survival horizon, if the difference represents true downstaging to stage III. Colorectal, lung, and esophageal cancers account for most of the projection. The estimate is a transparent scenario, not proof that MCED screening reduces mortality.

## Data Availability

data are available from the corresponding author upon reasonable request

